# Increased Autotaxin levels in severe COVID-19, correlating with IL-6 levels, endothelial dysfunction biomarkers, and impaired functions of dendritic cells

**DOI:** 10.1101/2021.07.30.21261361

**Authors:** Ioanna Nikitopoulou, Dionysios Fanidis, Konstantinos Ntatsoulis, Panagiotis Moulos, George Mpekoulis, Maria Evangelidou, Alice G. Vassiliou, Vasiliki Dimakopoulou, Edison Jahaj, Stamatios Tsipilis, Stylianos E. Orfanos, Ioanna Dimopoulou, Emmanouil Angelakis, Karolina Akinosoglou, Niki Vassilaki, Argyris Tzouvelekis, Anastasia Kotanidou, Vassilis Aidinis

## Abstract

Autotaxin (ATX; *ENPP2*) is a secreted lysophospholipase D catalysing the extracellular production of lysophosphatidic acid (LPA), a pleiotropic signalling phospholipid. Genetic and pharmacologic studies have previously established a pathologic role for ATX and LPA signalling in pulmonary injury, inflammation, and fibrosis. Here, increased *ENPP2* mRNA levels were detected in immune cells from nasopharyngeal swab samples of COVID-19 patients, and increased ATX serum levels were found in severe COVID-19 patients. ATX serum levels correlated with the corresponding increased serum levels of IL-6 and endothelial damage biomarkers, suggesting an interplay of the ATX/LPA axis with hyperinflammation and the associated vascular dysfunction in COVID-19. Accordingly, dexamethasone (Dex) treatment of mechanically ventilated patients reduced ATX levels, as shown in two independent cohorts, indicating that the therapeutic benefits of Dex include the suppression of ATX. Moreover, large scale analysis of multiple single cell RNAseq datasets revealed the expression landscape of *ENPP2* in COVID-19 and further suggested a role for ATX in the homeostasis of dendritic cells, that exhibit both numerical and functional deficits in COVID-19. Therefore, ATX has likely a multifunctional role in COVID-19 pathogenesis, worth of suggesting that its pharmacological targeting might represent an additional therapeutic option.

## Introduction

The leading symptom of COVID-19, beyond cough and fever, is hypoxemia, leading to dyspnea in severe cases, attributed to impaired lung mechanics and/or vasoconstriction (1, 2). Endothelial dysfunction is also a major characteristic of COVID-19 (3), shared with hypertension, diabetes and obesity, the most common comorbidities that are associated with poor prognosis (1, 2). The respiratory epithelial cell damage that follows viral infection and replication stimulate, depending on the underlying genetic and metabolic context, a hyperinflammatory state denominated “cytokine storm” (4). The excessive production of pro-inflammatory cytokines, such as TNF and IL-6, further induces endothelial damage and lung injury, and its more severe form, Acute Respiratory Distress syndrome (ARDS), that can result to respiratory and/or multi-organ failure and death (5).

A subset of surviving COVID-19 ARDS-like patients will develop a fibroproliferative response characterized by fibroblast accumulation and ECM deposition (6), also evident in *postmortem* histopathological analysis of the lungs of COVID-19 patients (7). Moreover, many discharged COVID-19 patients present with abnormally pulmonary architecture and functions (8-12), suggesting persisting fibrotic abnormalities, pending long-term follow up studies. Single-cell RNA sequencing (scRNAseq) analysis and transcriptional profiling indicated similarities in expression profiles between Idiopathic pulmonary fibrosis (IPF) and COVID-19 (13, 14), while CoV-2 infection has been suggested to stimulate the expression of major pro-fibrotic factors including TGFβ (15). *Vice versa*, patients with interstitial lung diseases (ILD) had an increased risk for severe COVID-19 and poor outcomes (ICU admittance, death) following CoV-2 infection (16-18).

Autotaxin (ATX; ENPP2) is a secreted lysophospholipase D that can be found in most biological fluids, including blood and bronchoalveolar lavage fluid (BALF), largely responsible for the extracellular production of lysophosphatidic acid (LPA), a growth factor-like signaling phospholipid. Increased ATX expression and LPA signaling has been reported in cancer as well as in chronic inflammatory diseases (19), including IPF (20, 21). Genetic and pharmacologic studies have further uncovered a therapeutic potential for ATX in IPF (20, 22-24), leading to phase III clinical trials (25). Given the associations of COVID-19 with pulmonary fibrosis, the pro-fibrotic properties of ATX, as well the many reported LPA effects on pulmonary cells and especially the vasculature (26), in this study we explored a possible association of ATX with COVID-19. In this context, we quantified *ENPP2* mRNA levels in nasopharyngeal swabs, ATX protein levels in the sera of two cohorts of COVID-19 patients, while we performed a large-scale analysis of recently published scRNAseq COVID-19 datasets.

## Materials and Methods

### Human patients and samples

All studies were performed in accordance with the Helsinki Declaration principles. All collected data were anonymized in standardized forms and informed consent was obtained from all individuals or patients’ next-of-kin for severe cases. All available patient personal, epidemiological, clinical, and experimental data are summarized in the corresponding cohorts’ tables (1, 2 and 3), including the appropriate protocol approvals.

### Enzyme-linked immunosorbent assay (ELISA)

ATX and IL-6 protein levels were quantified with dedicated ELISA kits according to the manufacturer’s instructions (R&D Systems Inc., Minneapolis, MN, USA, and Invitrogen, ThermoFisher Scientific, CA, USA respectively). Measurements were performed in a blinded fashion in triplicates using the Triturus automated analyser (Grifols, Barcelona, Spain). The presented results on ELISA quantification of soluble E-selectin (sE-sel) and P-selectin (sP-sel), ICAM and ANG2 in the same samples, has been reported previously (27).

### RNA extraction and Q-RT-PCR

Total RNA extraction from nasopharyngeal swab samples was performed using the MagNA Pure LC Total Nucleic Acid Isolation Kit using the MagNa Pure LC 2.0 automated nucleic acid purifier (Roche), and viral RNA was quantified with the LightMix Modular SARS-CoV-2 RdRP Kit and the LightCycler Multiplex RNA Virus Master kit (Roche). *ENPP2* mRNA levels were quantified with Q-RT-PCR using the SYBR Green-based Luna® Universal qPCR Master Mix (New England Biolabs)(*ENPP2:* forward: 5’-ACT CAT GAA GAT GCA CAC AGC -3’; reverse 5’-CGC TCT CAT ATG TAT GCA GG -3’; product length 131 bp). Normalization was performed with the house-keeping gene 14-3-3-zeta polypeptide (YWHAZ), and the relative quantification method 2^-ΔΔCt^ was utilised.

### Bulk/ single cell RNA-seq data analysis and mining

The available single cell RNA-seq object was mined for each one of the datasets (Table 1) using Seurat package v3 (28). Marker selection and DEA were performed using Wilcoxon Rank Sum test (FC>1.2; Bonferroni adj. p< 0.05). For identifying pDCs in the lung data set of (13), DCs – as initially marked– were isolated, and principal components were calculated post to variable genes identification and data scaling using default parameters. The 30 first principal components were used to construct a SNN graph, while clusters were defined with a resolution of 0.8. pDCs were identified using marker genes reported in the cell atlas of (29).

**Table 1.** Increased *ENPP2* mRNA expression in nasopharyngeal swabs from COVID-19 patients compared to healthy, non-infected controls. Nasopharyngeal swab (NS) samples were collected upon routine diagnosis from adult patients tested positive in SARS-CoV2 RNA PCR and showing no or mild COVID-19 clinical symptoms, including cough, sore throat, mild fever below 38oC and loss of smell (positive mild group) or being hospitalized in the intensive care unit (ICU) with severe/critical symptoms, such as respiratory failure, septic shock, acute thrombosis and multiorgan dysfunction (positive severe/critical group). The control group consisted of individuals with a negative SARS-CoV2 RNA PCR.

|  | Negative<br>Healthy | Positive<br>Mild | Positive<br>Severe/critical |
| --- | --- | --- | --- |
| Number of patients (n) | 21 | 21 | 21 |
| **ATX** ($2^{-\Delta\Delta C_t}$, mean ± SD) | 2.15 ± 1.37 | 5.38 ± 2.34**** | 5.76 ± 2.18**** |
| **Sex** |  |  |  |
| Male | 8 (38.09%) | 10 (47.6%) | 17 (80.95%) |
| Female | 13 (61.9%) | 9 (42.8%) | 4 (19.04%) |
| Not recorded | - | 2 (9.52%) | - |
| **Age** (years, mean ±SD) | 50.15 ± 20.86 | 37.78 ± 11.89 | 63.38 ± 17.23 |

Preprocessed read count matrices of (30) found <u>here</u> were analyzed using metaseqR2 package (31). More specifically, reads where EDASeq normalized, filtered using default parameters and then PANDORA algorithm was used to combine results of DESeq (32), DESeq2 (33), limma-voom (34), edgeR (35) and ABSSeq (36) methods. DEGs were defined using a FC>1.2 and FDR adj. meta p-value < 0.05.

### Statistical analysis

Statistical significance was assessed with the Prism (GraphPad) software with the appropriate test according to the distribution of values and their complexity, as detailed in each figure legend. Statistical tests used include the non-parametric Mann-Whitney U test, unpaired t-test, Spearman corelation, 2-way ANOVA followed by Bonferroni post hoc correction, Wilcoxon rank sum test-Bonferroni correction, and Kruskal-Wallis and Dunn post hoc test.

## Results

### Increased *ENPPp2* mRNA levels in nasopharyngeal swab samples from COVID-19 patients

As viral infections have been reported to stimulate *ENPP2* mRNA expression (37, 38), and to examine if CoV-2 infection has similar effects, we first quantified *ENPP2* mRNA levels with Q-RT-PCR in nasopharyngeal swab samples (Table 1). A significant increase was found in *ENPP2* mRNA expression in mild and severe COVID-19 patients, as compared to non-infected subjects (Fig. 1). Therefore, CoV-2 infection stimulates *ENPP2* mRNA expression in the respiratory epithelial or immune cells that compose the nasopharyngeal swab samples.

**Figure 1.**
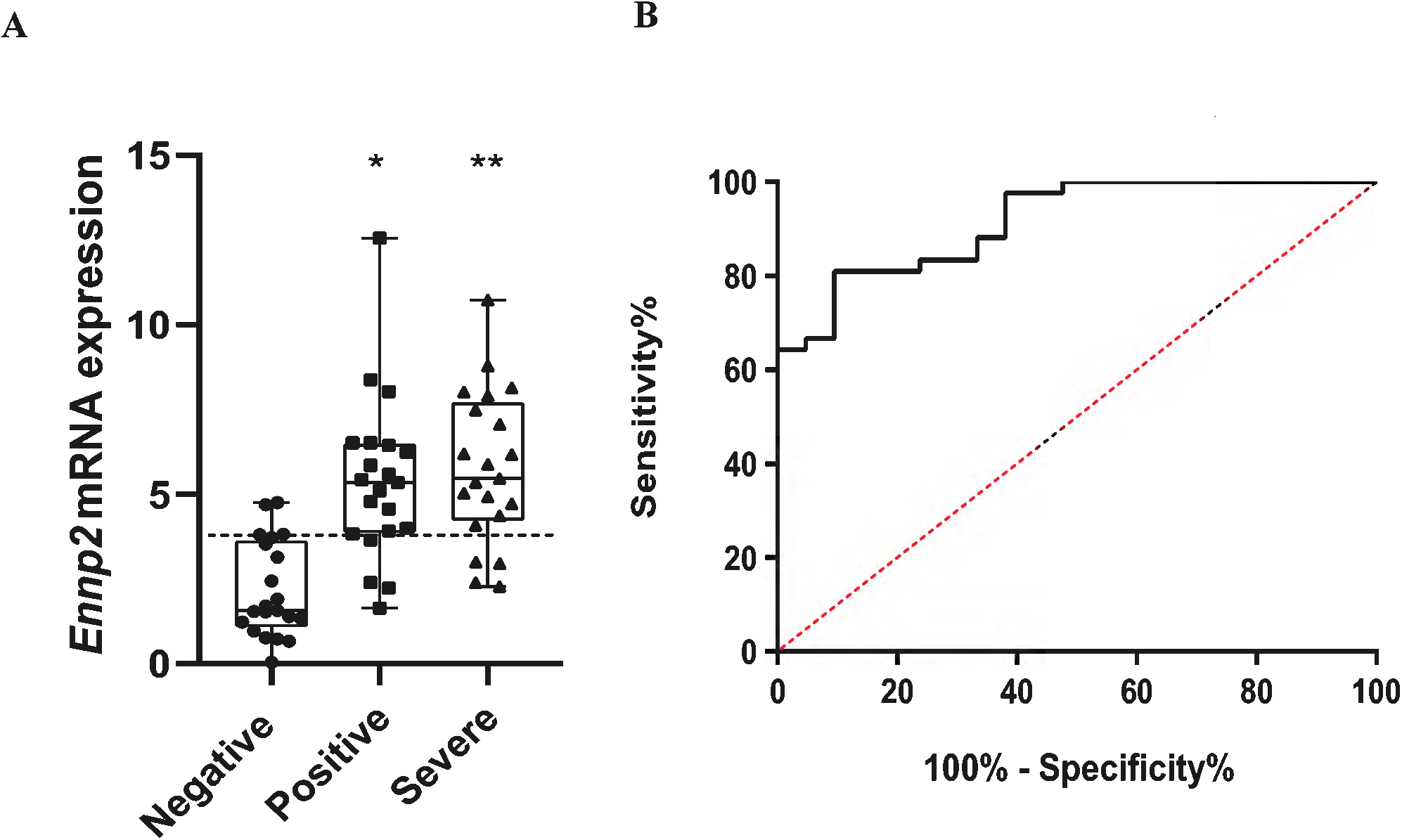
Increased *ENPP2* mRNA expression in nasopharyngeal swab samples from patients with mild or severe/critical COVID-19. **(A)**. *ENP2* mRNA values (mean 2-M^ct^) from the two groups of patients (n=21 per group) and the control group (n=21). The horizontal dotted line indicates the optimal threshold value (cut-off). Data are represented as box plots; line in the middle, median; box edges, 25^th^ to 75^th^ centiles; whiskers, range of values. p values were calculated with the non-parametric Mann-Whitney U test. **(B)**. ROC curves were generated after merging the results for the two positive groups (mild and severe/critical) and AUC, 95% CI, p values and cut-off points with their specificity and sensitivity were calculated (table). Positive mild versus negative samples: * p= 4⨯10^-6^, Positive critical/severe versus negative samples: ** p= 2.92⨯10^-7^.

### Increased serum ATX protein levels in severe COVID-19 patients

To examine if systemic levels of ATX are possibly increased upon COVID-19, ATX was quantified with an ELISA kit in the serum of COVID-19 patients hospitalised at the Evangelismos University Hospital (Table 2). The cohort consisted of patients with mild symptoms, hospitalised in the COVID-19 WARD (n=47; no Dex treatment), as well as of patients with severe symptoms, hospitalised in the Intensive Care Unit (ICU); ICU patients were further separated in patients receiving Dexamethazone (Dex) treatment (n=37) or not (NO Dex; n=32). A large proportion of patients suffered from comorbidities, and were receiving a variety of medications prior to admission, while COVID-19-targeted treatment included azithromycin, chloroquine and lopinavir/ritonavir in different combinations per WHO recommendations at that time (Table 2). In comparison with WARD patients, ICU patients were hypoxemic (low ratio of arterial oxygen partial pressure to fractional inspired oxygen; PaO2/FiO2), lymphopenic (low lymphocyte numbers), and had increased LDH levels (Table 2), all three suggested as disease severity markers.

**Table 2.** Clinical characteristics and laboratory data of COVID-19 patients hospitalized at the Evangelismos general hospital. Serum samples were collected with standardised procedures from COVID-19 patients admitted to the specialized COVID-19 WARD or to the intensive care unit (ICU) of the Evangelismos General Hospital from 24 March to 2 November 2020. SARS-CoV-2 infection was diagnosed by real-time reverse transcription PCR in nasopharyngeal swabs. The study was approved by the Evangelismos Hospital Research Ethics Committee (# 170/24-4-2020).

|  | WARD no Dex | ICU no Dex | ICU + Dex |
| --- | --- | --- | --- |
| Number of patients (n) | 47 | 37 | 32 |
| ATX (ng/ml, mean ±SD) | 310.32 ± 98.85* | **443 ± 172.90** | 246.15 ± 73.74* |
| **Sex** |  |  |  |
| Male | 33 (70.21%) | 31 (83.78%) | 22 (62.5%) |
| Female | 14 (29.78%) | 6 (16.21%) | 10 (31.25%) |
| **Age** (years, mean ±SD) | 54.63 ± 15.46 | 63.54 ± 10.89 | 65.5 ± 10.7 |
| **Comorbidities** n (%) |  |  |  |
| Hypertension | 13 (27,65%) | 17 (45.94%) | 12 (37.5%) |
| Diabetes | 4 (8,51%) | 5 (13.51%) | 5 (15.62%) |
| Coronary artery disease | 8 (17,02%) | 4 (10.81%) | 4 (12.5%) |
| COPD | 1 (2,12%) | 1 (2.7%) | 2 (6.25%) |
| Asthma | 2 (4,25%) | 1 (2.7%) | 1 (3.12%) |
| Hyperlipidaemia | 9 (19,14%) | 9(24.32%) | 8 (25%) |
| Hepatitis | 0 (0%) | 1 (2.7 %) | 0 (0%) |
| **COVID-19 treatment** |  |  |  |
| Azithromycin/chloroquine/lopinavir/ritonavir | 0 | 11 |  |
| Azithromycin/chloroquine | 6 | 7 |  |
| Lopinavir/ritonavir/chloroquine | 0 | 2 |  |
| Chloroquine | 0 | 3 |  |
| Plasma | 0 | 1 |  |
| **Clinical measurements** |  |  |  |
| Mean arterial pressure (mmHg) | 83,19 ± 8.86 | 82.83 ± 16.52 | 77.55 ± 8.54 |
| PaO<sub>2</sub>/FiO<sub>2</sub> (mmHg) | 301.5 ± 79.81* | **194.86 ± 86.64** | 85.94 ± 15.97* |
| Glucose (mg/dL) | 133,5 ± 113,3 | 164.53 ± 77.73 | 164.06 ± 75.40 |
| Creatinine (mg/dL) | 0.9 ± 0.33 | 1.02 ± 0.32 | 0.95 ± 0.72 |
| CRP (mg/dL) | 6,8 ± 8,96 | 14 ± 10.17 | 13.83 ± 9.6 |
| Total bilirubin (mg/dL) | 0.5 ± 0.33 | 0.73 ± 0.5 | 0.61 ± 0.29 |
| White blood cell count (per µl) | 6995 ± 3468 | 10125 ± 4633 | 11,705 ± 10,372 |
| Neutrophils (%) | 69,34 ± 13,51 | 81.34 ± 6.64 | 83.12 ± 12.2 |
| Lymphocytes (%) | 24,03 ± 10,89* | **12.63 ± 5.63** | 11.12 ± 11.23 |
| Platelets (per µl) | 240297 ± 110028 | 237783 ± 101338 | 257000 ± 79581 |
| INR (median IQR) | 1.06 ± 0.09 | 2.07 ± 5.73 | 1.26 ± 0.65 |
| D-dimer (pg/ml) | 1.19 ± 1.72 | 0.47 ± 0.26 | 1.39 ± 0.93 |
| AST (IU/L) | 36,65 ± 30,65 | 54.18 ± 39.95 | 121.4 ± 329.9 |
| ALT (IU/L) | 33,15 ± 23,58 | 45.9 ± 28.08 | 60.8 ± 72.4 |
| LDH (U/L) | 286,36 ± 122,08* | **498.48 ± 242.34** | 591.23 ± 490.84 |
| Fibrinogen (mg/dL) | 514,06 ± 176,18 | 638.18 ± 158.76 | 630.3 ± 172.2 |
| Ferritin (pg/ml) | 513,48 ± 815,55 | 2786 ± 694.48 | 912.47 ± 826.91 |
| APACHE II score | 5.25 ± 2.94 | 14.27 ± 5.08 | 15.4 ± 3.89 |
| SOFA score | 2 ± 1 | 6.83 ± 3.08 | 5.4 ± 1.81 |

Increased ATX serum concentrations were discovered in ICU patients (not receiving Dex) as compared with WARD patients (Fig. 2A), suggesting a possible association of ATX with disease severity. However, no substantial, statistically significant corelation was observed independently with the applicable severity markers (data not shown and Table 2); no statistically significant differences of ATX levels between the sex or the comorbidities of COVID-19 patients was detected either (Fig. S1). However and most importantly, ATX levels corelated significantly with IL-6 levels in the serum of ICU patients (not receiving Dex)(Fig. 2B), suggesting a possible interplay of ATX/LPA with the cytokine storm in COVID-19.

**Figure 2.**
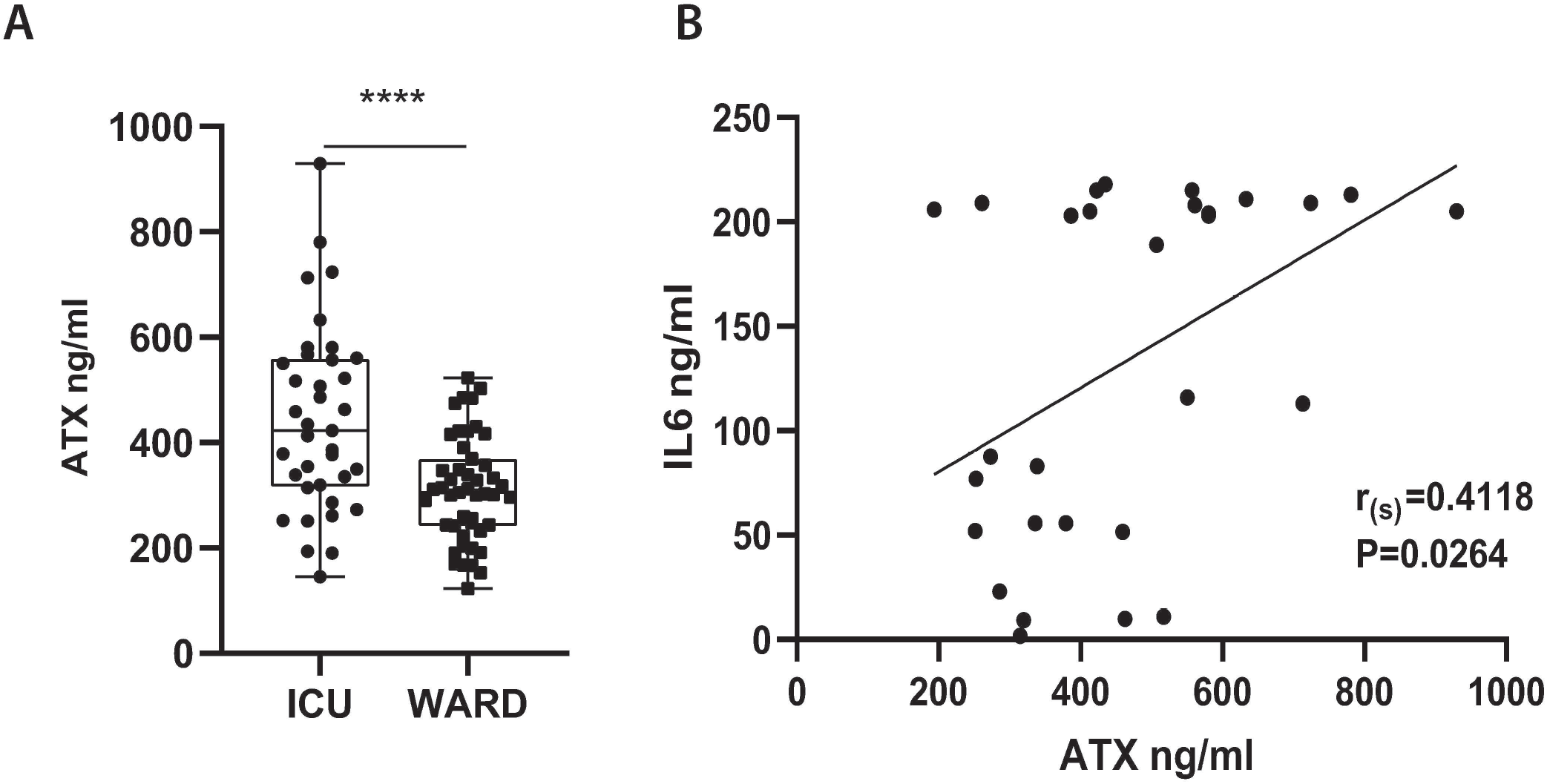
Increased serum ATX protein levels in COVID-19 patients hospitalized in the intensive care unit (ICU), corelating with increased IL-6 levels. **(A)**. ATX protein levels were measured, with a commercial ELISA kit, in the sera of COVID-19 patients hospitalized (without Dex treatment) in the COVID-19 Ward (n=47) or the ICU (n=37) of Evangelismos hospital. Statistical significance, given the normal distribution of values, was assessed with an unpaired t-test. ****denotes p<0,0001. (B). ATX serum levels correlated with serum IL-6 levels (n=29), as assessed with Spearman correlation (r_s_).

ICU non-survivors in this cohort had higher levels of the endothelial dysfunction markers soluble E-selectin (sE-sel), soluble P-selectin (sP-sel), soluble intercellular adhesion molecule 1 (sICAM-1) and angiopoietin 2 (ANG-2) when compared to survivors, as recently reported using a subset of the current Evangelismos cohort samples (27). Interestingly, the increased ATX protein levels correlated with the increased protein levels of sEsel and sICAM (Fig. S2) in ICU patients, suggesting a role for ATX/LPA in COVID-19 induced endothelial dysfunction.

### Dex therapeutic effects in COVID-19 include the suppression of ATX serum levels

The first line of therapy for many inflammatory diseases as well as respiratory infections is Dex, that lowers the expression of pro-inflammatory cytokines including IL-6, and that has been proven effective in COVID-19 patients requiring, invasive or not, oxygenation (39, 40). Therefore, we next examined ATX serum levels in intubated, or not, ICU patients receiving, or not, Dex treatment. Remarkably, Dex treatment was discovered to potently suppress ATX serum levels in ventilated patients (Fig. 3A), while intubated ICU patients receiving no Dex presented with the highest overall ATX serum levels. Identical results were obtained in another cohort of ICU patients from the University hospital of Patras (Table 3)(Fig. 3B), indicating that the therapeutic benefits of Dex include the suppression of ATX serum levels.

**Table 3.** Clinical characteristics and laboratory data of COVID-19 patients hospitalized at the ICU of the University Hospital of Patras. Serum samples were collected with standardised procedures from COVID-19 patients admitted to intensive care unit (ICU) of the University Hospital of Patras from April 24^th^ to December 6^th^ 2020. The study was approved by the University Hospital of Patras Research Ethics Committee (# 216/08-05-2020).

|  | ICU no Dex | ICU + Dex |
| --- | --- | --- |
| Number of patients (n) | 12 | 23 |
| ATX (ng/ml, mean ±SD) | 624.36 ± 203.5 | **404.16 ± 145.5\*\*** |
| **Sex** |  |  |
| Male | 9 (75%) | 18 (78.26%) |
| Female | 3 (25%) | 5 (21.73%) |
| Age (years, mean ±SD) | 66.75 ± 13.31 | 59.43 ± 15.42 |
| **Comorbidities n (%)** |  |  |
| Hypertension | 5 (41.6%) | 10 (43.47%) |
| Diabetes | 0 (0%) | 4 (17.39%) |
| Coronary artery disease | 2 (16.6%) | 1 (4.34%) |
| COPD | 0 (0%) | 2 (8.69%) |
| Asthma | 0 (0%) | 0 (0%) |
| Hyperlipidaemia | 3 (25%) | 6 (26.08%) |
| Hepatitis | (%) | (%) |
| **COVID 19 treatment** |  |  |
| Azithromycin/chloroquine/lopinavir/ritonavir | 4 (33.33%) | 0 (0%) |
| Azithromycin/chloroquine | 6 (50%) | 5 (21.73%) |
| Lopinavir/ritonavir/chloroquine | 1 (8.33%) | 0 (0%) |
| Chloroquine | 0 (0%) | 0 (0%) |
| Plasma | (%) | (%) |
| **Clinical measurements** |  |  |
| Glucose (mg/dL) | 120.5 ± 24.57 | **154.04 ± 47.98\*** |
| Creatinine (mg/dL) | 0.95 ± 0.42 | 0.93 ± 0.38 |
| CRP (mg/dL) | 8.53 ± 5.51 | 15.55 ± 12.68 |
| Total bilirubin (mg/dL) | 0.67 ± 0.27 | 0.7 ± 0.36 |
| Lymphocytes (absolute number) | 0.62 ± 0.35 | 0.83 ± 0.5 |
| INR (median IQR) | 1.09 ± 0.13 | 1.07 ± 0.12 |
| D-dimer (pg/ml) | 2.21 ± 2.14 | 1.45 ± 1.72 |
| LDH (U/L) | 370 ± 129 | 455.08 ± 188.56 |
| Fibrinogen (mg/dL) | 630.55 ± 168.1 | 545 ± 182.32 |
| Ferritin (pg/ml) | 950 ± 382.64 | 1131.09 ± 1223.55 |

*Statistical significance was assessed with an unpaired t-test; \*, \*\*denotes $p<0.05$ , $p<0.01$ . ATX values appear at Figure 3.*

**Figure 3.**
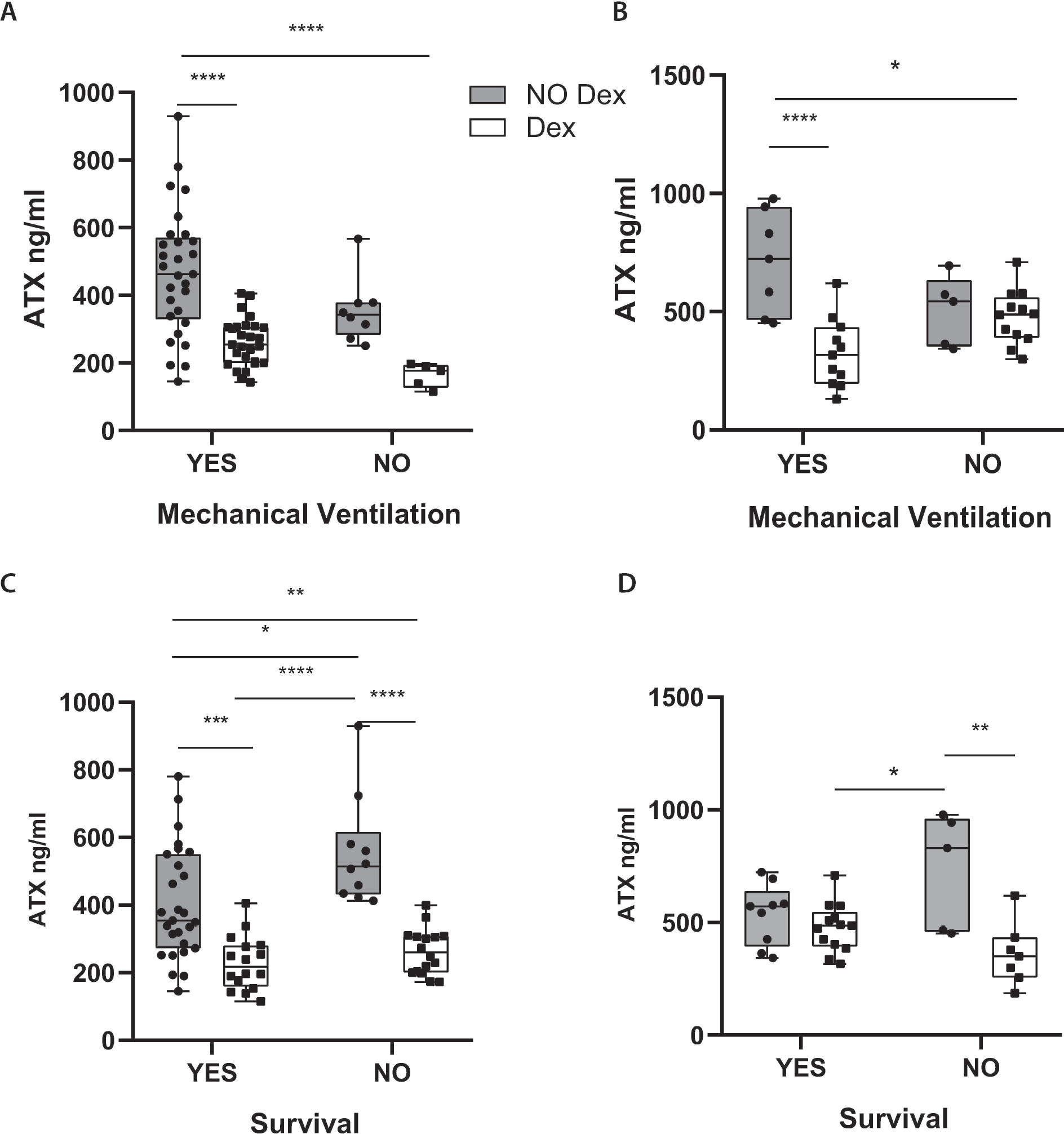
Dexamethasone therapeutic effects include the suppression of ATX serum levels. ATX protein levels were measured, with a commercial ELISA kit in the serum of COVID-19 patients hospitalized in the ICU of **(A, C)** the Evangelismos or **(B, D)** the Patras hospitals. The measurements in the Dex groups in 3A and C, are the same as in Figure 2A. Statistical significance, given the normal distribution of values, was assessed with 2-way ANOVA followed by Bonferroni post hoc correction. *, **, ***, **** denotes p<0.05, p< 0.01, p<0.001 and p<0.0001 respectively.

Moreover, ATX levels in ICU patients not receiving Dex treatment negatively affected survival, and non-surviving ICU patients receiving no Dex presented with the higher overall ATX serum levels (Fig. 3 C,D).

### The *ENPP2* expression landscape in COVID-19

To identify possible ATX expressing cells in the nasopharyngeal swab (NS) samples (Fig. 1), peripheral blood monocytes (PBMCs) in the circulation (Figs. 2, 3), as well as in BALF and lung tissue cells, we re-analysed and mined several scRNAseq datasets of COVID-19 patients and healthy controls, from recent high impact studies (Table 1), collectively interrogating the gene expression of more than 10^6^ cells; cell clustering and naming followed that of the original analyses, which both varied between studies/datasets.

In NS cells, ATX is mainly expressed by natural killer cells (NKs) and monocyte-derived macrophages (MoAM)(Figs. 4A and S3A), as detected in two NS datasets (Table 1). In the circulation, and in both PBMCs datasets (Table 1), *ENPP2* expression was mainly detected, surprisingly, in plasmacytoid DCs (pDCs; Figs 4B and S3B). In BALF cells (Table 1), *ENPP2* expression was also mainly detected in pDCs, as well as MoAMs (Figs 4C and S3C). In lung tissue (Table 1), *ENPP2* was found to be primarily expressed in arterial and mesothelial cells, as well as in cells of the monocytic lineage (Fig. 4D and S3D). A similar lung tissue profile was also detected (Fig. S3E) in an IPF scRNAseq dataset (Table 1), extending the similarities of pathogenic mechanisms between IPF and COVID-19 and supporting a common role for ATX.

**Figure 4.**
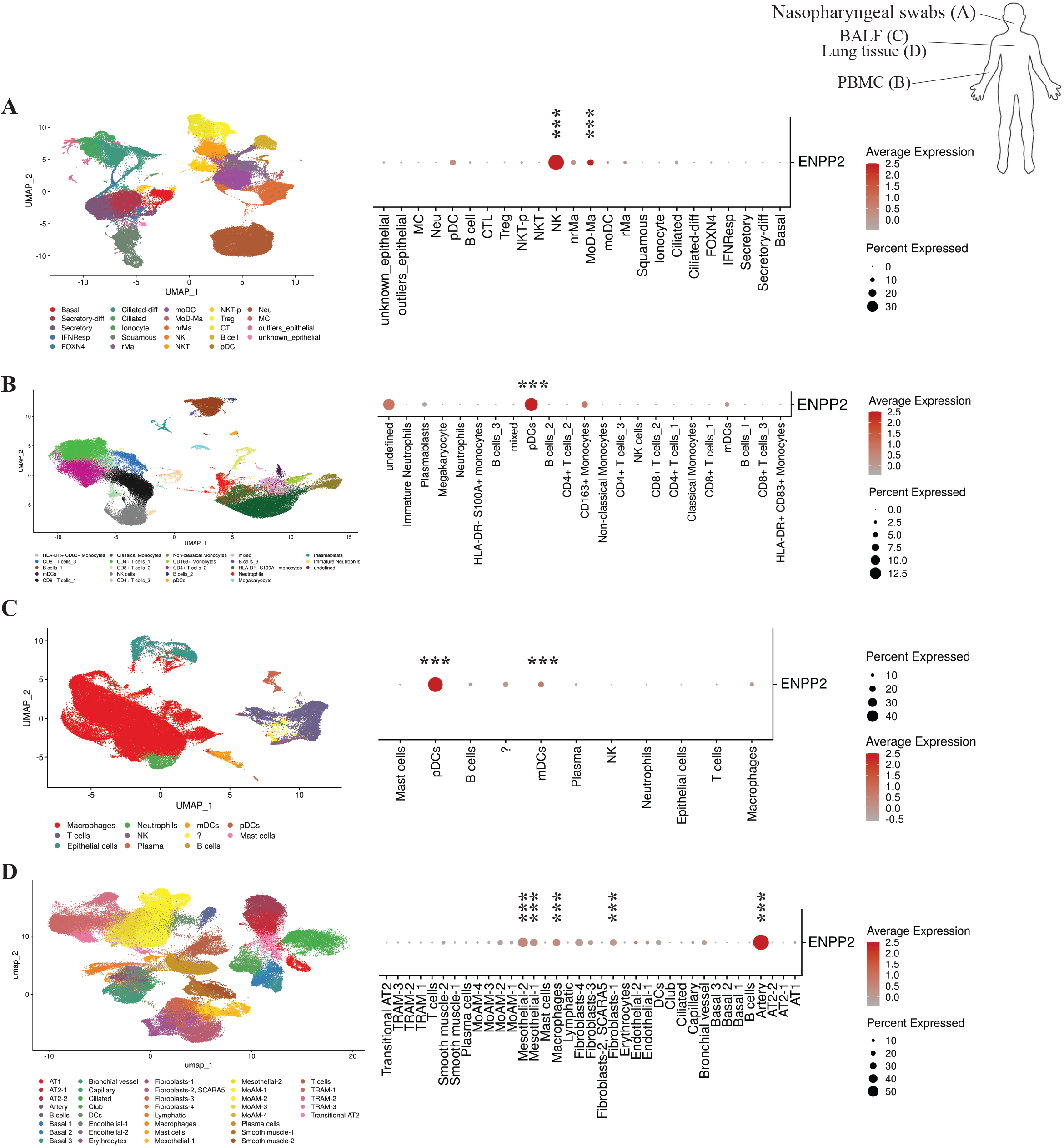
The *ENPP2* expression landscape in COVID-19. *ENPP2* expression has been assessed in four datasets of COVID-19/healthy control datasets, each representing a different sampling site (Table 1). UMAP plots (on the left) depict the cellular composition of these sites, while dot plots (on the right) the expression pattern of *ENPP2* in the detected cell types. Marker genes, denoted by stars, were detected using a Wilcoxon rank sum test; FC>1.2, Bonferroni corrected p<0.05; ^***^denotes p< 0.01) (PMIDs: A. <u>32591762</u>; B. <u>32810438</u>; C. <u>32398875</u>; D. <u>33257409</u>).

### A role for ATX in the homeostasis of dendritic cells

Given the *ENPP2* expression from monocytic cells and especially pDCs, we next interrogated *ENNP2* mRNA levels specifically in pDCs from COVID-19 patients in comparison with control samples, subsets of the datasets analysed in Figure 4. Confined by the limited numbers of lung pDCs, as well as the detected genes per cell and the relatively low expression levels of *ENPP2*, the analysis indicated a statistically significant overexpression of *ENPP2* in COVID-19 circulating pDCs (Fig. 5B). Noteworthy, DCs are the highest *ENPP2* expressing immune cells during healthy conditions, as identified upon querying a large-scale RNAseq data set interrogating gene expression of 28 immune cell types (79 healthy volunteers and 337 patients from 10 immune-related diseases)(30)(Fig. S4A). Similar analysis indicated that the main LPA receptor expressed by DCs is LPAR2 (Fig. S4B), which has been suggested to convey anti-inflammatory LPA signals to DCs (41). Furthermore, increased *ENPP2* mRNA expression was detected in pDCs from patients with systemic lupus erythematosous (SLE), adult-onset Still’s disease (AOSD), mixed connective tissue disease (MCTD) and idiopathic inflammatory myopathy (IIM) than in DCs from healthy volunteers (Fig. S4C), suggesting that overexpression of *ENPP2* in pDCs may be a common theme in inflammation.

**Figure 5.**
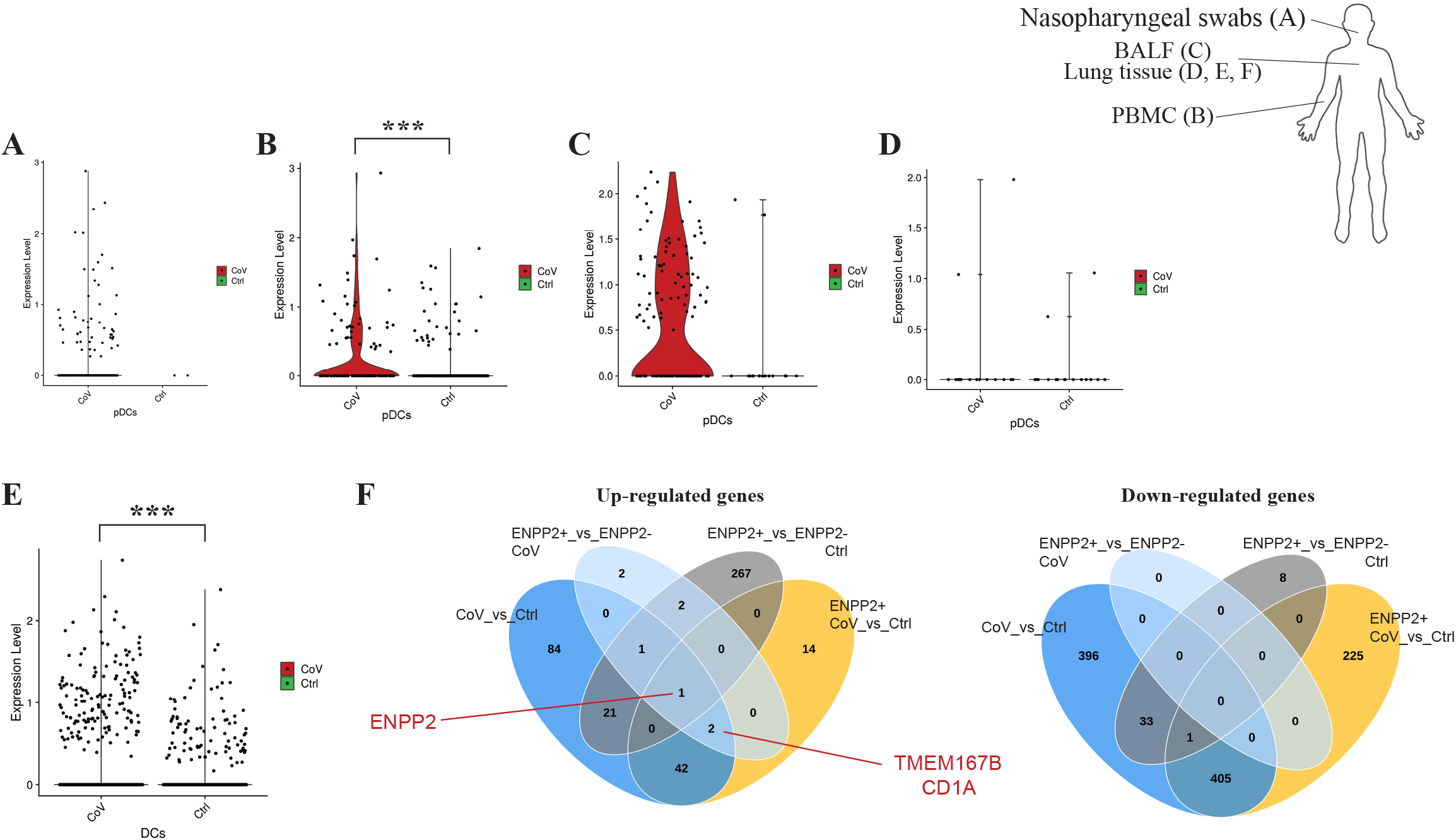
*ENPP2* mRNA expression is upregulated in peripheral pDCs and lung tissue DCs of COVID-19 patients. **(A-D)**. Differential expression of *ENPP2* in pDCs of COVID-19 patients versus healthy controls. *ENPP2* mRNA expression was found up-regulated in peripheral (B) pDCs of COVID-19 patients. **(E)**. Differential expression analysis indicates increased *ENPP2* mRNA expression in COVID-19 lung tissue DCs than healthy control ones. **(F)**. Venn diagrams of deregulated genes in lung DCs. Differential expression was performed using a Wilcoxon rank sum test; FC>1.2 and Bonferroni corrected p<0.05; ^***^denotes Bonferroni adjusted p< 0.01) (PMIDs: A. <u>32591762</u>; B. <u>32810438</u>; C. <u>32398875</u>; D. <u>33257409</u>).

Finally, and to gain mechanistic insights into the possible role of ATX in DCs homeostasis upon COVID-19, we first analysed differential gene expression in COVID-19 DCs (as pDCs were too few), from the only COVID-19 lung dataset (13) allowing such analysis, as well as in *ENPP2*-expressing (*ENPP2*^+^) DCs (Table S5). Increased *ENPP2* expression was also detected in all lung DCs (Fig. 5E), while comparative analysis (Venn diagrams Fig. 5F) highlighted two genes upregulated in *ENPP2*^+^ COVID-19 DCs, transmembrane protein 176B (TMEM176B) and CD1a, that have been both proposed as DC differentiation and/or maturation markers, suggesting that *ENPP2* expression may modulate DC homeostasis.

## Discussion

Previous studies have shown that HCV, HIV and HBV viral increase *Enpp2* mRNA expression in infected cells and/or to raise systemic ATX levels (42-44). As shown here, increased *ENPP2* mRNA expression was detected in nasopharyngeal swab samples from COVID-19 patients in comparison to non-infected healthy controls (Fig. 1), while scRNAseq re-analysis revealed that the highest *ENPP2* expressing cells in swabs are immune cells (Figs. 5A and S5A), suggesting that CoV-2 infection stimulates *ENPP2* expression from immune cells in the nasopharynx. LPA, the enzymatic product of ATX and its effector molecule, has been shown to directly affect HCV viral infection and replication (37, 38), suggesting that a similar autocrine mode of action maybe in play in COVID-19, where ATX produced by the infected host cell would stimulate local LPA production, in turn facilitating viral entry and replication.

Increased serum ATX protein have been reported in cancer, liver diseases, as well as respiratory diseases including asthma and pulmonary fibrosis (19, 45), while increased levels of serum ATX were recently reported in ARDS (46). Here, increased ATX sera levels were detected in ICU-hospitalised COVID-19 patients (receiving no Dex treatment) compared to patients with less severe disease (Fig. 2), suggesting increased ATX expression as an additional commonality of ARDS and COVID-19.

The origin of serum ATX is not completely deciphered; however, >40% of serum constitutively active ATX has been suggested to originate from the adipose tissue (47), which was shown to be able to modulate the pathophysiology of distant metabolically active organs (48, 49). Moreover, serum ATX has been reported to correlate with insulin resistance in older humans with obesity (50), while mice with heterozygous Enpp2 deficiency were protected from HFD-induced obesity and systemic insulin resistance (49). Several additional reports have incriminated the ATX/LPA axis in the regulation of glucose homeostasis and insulin resistance (reviewed in (51)), among the main comorbidities of COVID-19, suggesting adipose tissue-derived ATX as a possible pathologic link between obesity and COVID-19.

An additional possible source of serum ATX in disease states, beyond the adipose tissue, is the liver. Increased ATX expression has been reported in chronic liver diseases of different aetiologies, associated with shorter overall survival (37), while the genetic deletion of ATX from hepatocytes (37), or as discussed above adipocytes (48), attenuated liver steatosis and fibrosis. Therefore, increased levels of serum ATX are expected upon liver damage, whereas aberrant liver functions have been reported in COVID-19, irrespectively of pre-existing liver disease (52). On the other hand, cirrhotic patients have high rates of liver failure and death from respiratory failure upon CoV-2 infection, attributed to increased systemic inflammation, immune dysfunction, and vasculopathy (52). Therefore, ATX could be also a pathologic link between liver damage and COVID-19.

Plasma ATX levels have been recently reported to corelate with IL-6 levels in severe ARDS patients (46), as well as acute-on-chronic liver failure (ACLF) patients (53), as shown here in the serum of ICU COVID-19 patients (Fig. 2). Increased serum IL-6 levels have been reported in COVID-19 patients corelating with the severity of COVID-19 pneumonia and mortality risk (54), or respiratory failure and the need for mechanical ventilation (55). Meta-analyses of published studies on COVID-19 laboratory findings indicated that serum levels of IL-6 were among the most predictive biomarkers (56, 57). Interestingly, components of the COVID-19 cytokine storm (IL-6, TNF and IL-1β) have been suggested to stimulate ATX expression and/or activity in different cell types, while, *vice versa*, LPA has been reported to stimulate TNF and IL-6 expression in different contexts (22), suggesting a possible interplay of the COVID-19 cytokine storm and the ATX/LPA axis.

Dex treatment, a potent suppressor of systemic inflammation including IL-6, has been shown to reduce mortality in hospitalised COVID-19 patients under oxygen supplementation treatment or mechanical ventilation (39, 40). Dex treatment has been shown to decrease ATX (as well as IL-6) levels in the mouse adipose tissue and plasma (58), as well as in irradiated mammary fat pad (59). As shown here (Fig. 3), Dex treatment of mechanically ventilated patients drastically reduced their ATX serum levels, indicating that the therapeutic effects of Dex in COVID-19 include the suppression of ATX serum levels.

An essential role for ATX/LPA in embryonic vasculature has been well established through genetic studies in both mice (60-62) and zebrafish (63). In adult mice, *Enpp2* has been discovered as a high priority candidate gene for pulmonary hemorrhage upon SARS/MERS-CoV infection (64, 65), while vascular leak has been suggested to be among the main pathological effects of ATX/LPA in pulmonary pathophysiology and fibrosis in mice (21, 22). As shown here, *ENPP2* mRNA expression in the COVID-19 lung tissue was detected mainly in artery cells (Fig. 4D, S3D), while high ATX expression from ECs in HEVs in lymph nodes has been previously reported (66). Moreover, and in the same context, a plethora of LPA *in vitro* effects on endothelial cells have been suggested, with some controversy, including endothelial permeability, leukocyte adhesion, and cytokine expression, as previously reported in detail (26). Among them, LPA has been suggested to stimulate the expression of E-Sel from human aortic endothelial cells (67-69), a cell surface adhesion molecule regulating interaction with leukocytes. As shown here, ATX serum levels corelated with the corresponding sE-sel and sICAM serum levels (Fig. S2), which has been independently associated, in the same samples, with mortality of COVID-19 ICU patients (27), suggesting that ATX/LPA effects in COVID-19 may also include vasculopathy.

IPF macrophages have been previously shown to stain for ATX, and conditional genetic deletion of ATX from macrophages in mice, reduced BALF ATX levels and disease severity in modeled pulmonary fibrosis (20). scRNAseq analysis of BALF cells from COVID-19 patients, where macrophages predominate, indicated that *ENPP2* mRNA expression was detected in different macrophage subpopulations (Fig. 4C/UMAP, S3C/UMAP), where it could modulate their maturation in an autocrine mode via LPA (70-72). LPA has been also suggested to stimulate, *in vitro*, the conversion of monocytes to DCs (41, 73, 74). Interestingly, *ENPP2* mRNA expression was mainly detected in pDCs among all PBMCs and BALF cells in COVID-19 (Figs. 4B,C, S3B,C). pDCs are the principal interferon (IFN) type I producing cells in the human blood and can be rapidly recruited to inflamed sites (75). Circulating and lung pDCs have been shown to be diminished in COVID-19 (76, 77), while IFN type I responses were highly impaired (78, 79). *ENPP2* mRNA expression was found upregulated in circulating pDCs (Fig. 5B), and lung DCs (Fig. 5E) from COVID-19 patients in comparison to cells from healthy controls. pDC development and homeostasis are regulated by the transcription TCF4 (80), which has been reported to get modulated by LPA in colon cancer cells (81), suggesting that *ENPP2* expression from pDCs and the local production of LPA modulates, in an autocrine way, pDC development and homeostasis. The hypothesis is further supported from the genes that have been discovered, to be increased in COVID-19 DCs, possibly regulated by *ENPP2* (Fig. 5E). CD1a binds and presents to T-cells lipid metabolites and PLA2-synthesised fatty acid neoantigens and has been found to be expressed in immature DCs in mucosal surfaces, including the bronchus (82-84). Tmem176B has been also associated with an immature state of dendritic cells (85, 86), suggesting that *ENPP2* expression from COVID-19 pDCs, via LPA, delays their maturation. Although LPA signals in most cell types are considered pro-inflammatory and pro-surviving, an anti-inflammatory role of LPA, via LPAR2 -the main subtype expressed in DCs (Fig. S4B), has been proposed previously for DCs (41), further supporting a possible role for ATX/LPA in supressing DC responses.

Taken together, a role for ATX/LPA in COVID-19 pathogenesis seems likely, possibly as a component of the cytokine storm perpetuating hyperinflammation and stimulating endothelial damage, as well as a regulator of the mononuclear phagocyte system and a suppressor of (p)DCs responses, non-withstanding its established role in fibrosis. Dex treatment in mechanically ventilated patients decreased ATX levels, indicating that the therapeutic effects of Dex in COVID-19 include the suppression of the ATX/LPA axis and that ATX levels can be druggable. Additional large-scale studies of serum ATX levels, e.g. retrospectively from recent clinical trials (a-IL-6R, Dex, antiviral), will be required to possibly establish ATX as a diagnostic/prognostic marker. Moreover, perspective FACS analysis of BALF DCs from COVID-19 patients will be necessary to validate the scRNAseq -based hypotheses data on DC homeostasis. More importantly and given that COVID-19 and IPF share risk factors for disease severity, such as age/sex and comorbidities, existing and developing anti-fibrotic therapies have been suggested as additional therapeutic opportunities in COVID-19 (87-89). One of the novel candidates target ATX, is currently in clinical trials phase III in IPF (90). Given the multiple possible roles of ATX in COVID-19, ATX inhibition could offer additional therapeutic options in COVID-19 management, both during and after hospitalization.

## Supporting information

All Sup Figures

## Data Availability

All data are reported within the manuscript.

## Acknowledgements

We would like to thank Alexandros Zacharis and Nikolaos Athanasiou for sample collection.

## Author contributions

IN performed all ELISAs. IN, KN and PM performed all related statistical analyses. DF analyzed bulk and sc-RNAseq data. GM and ME performed Q-RT-PCRs on nasopharyngeal swabs and analyzed data with EA and NV. AV, VD, EJ and ST collected clinical samples and compiled clinical data under the supervision and guidance of SO, ID and KA. NV, AT, AK and VA designed the study and supervised analyses. VA wrote the paper, which was edited and critically commented by all authors, that all approved the reported conclusions.

---

**Table S1.**
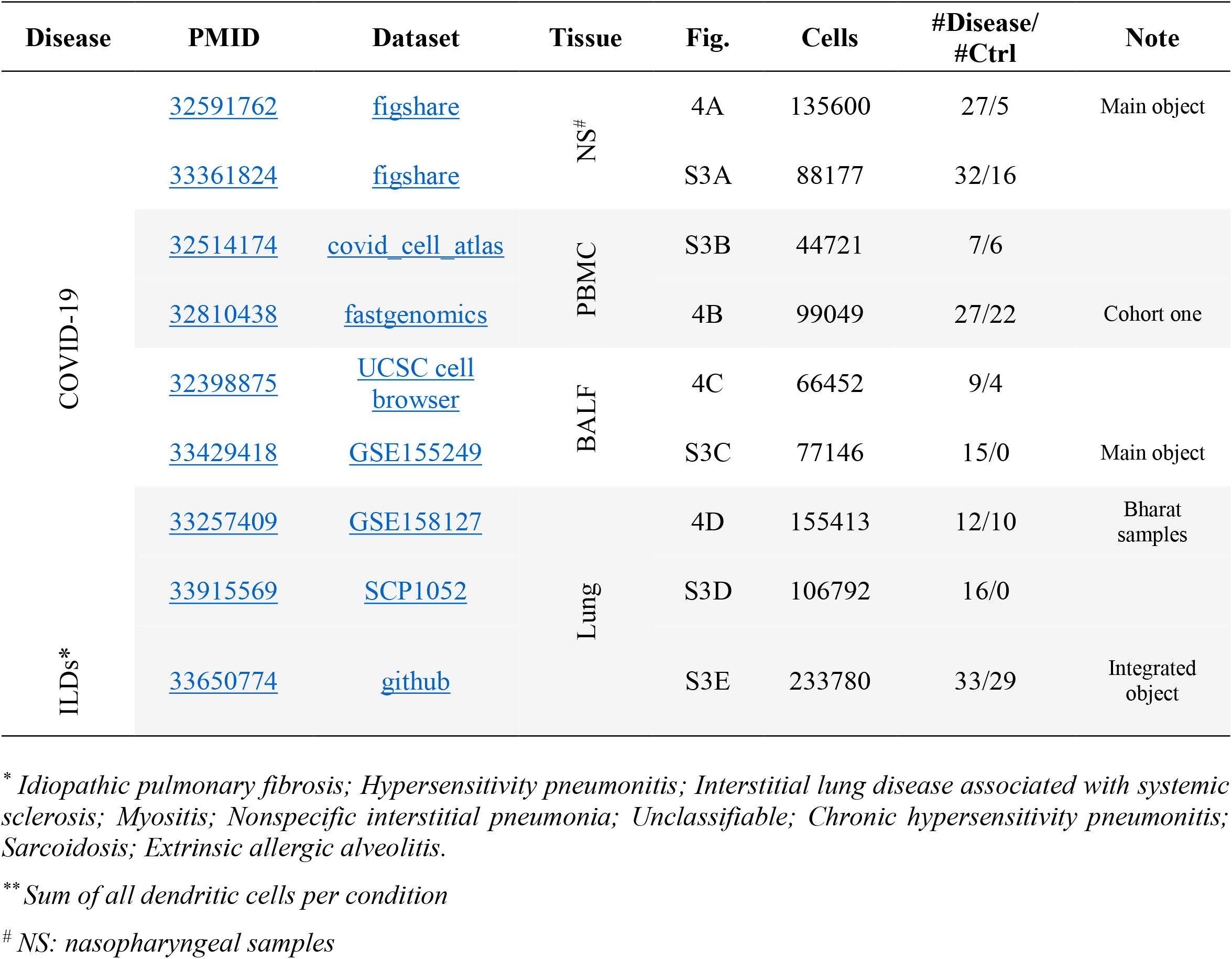
scRNA-seq datasets used in the study

**Table S2. Differential expression analysis of dendritic cells (DCs) in COVID-19**.

The corresponding spreadsheets can be found at: https://www.dropbox.com/s/zdq0z0ikxrraj37/Table%20S2.xlsx?dl=0

