## Supplementary material for "Increased Autotaxin levels in severe COVID-19, correlating with IL-6 levels, endothelial dysfunction biomarkers, and impaired functions of dendritic cells": All Sup Figures

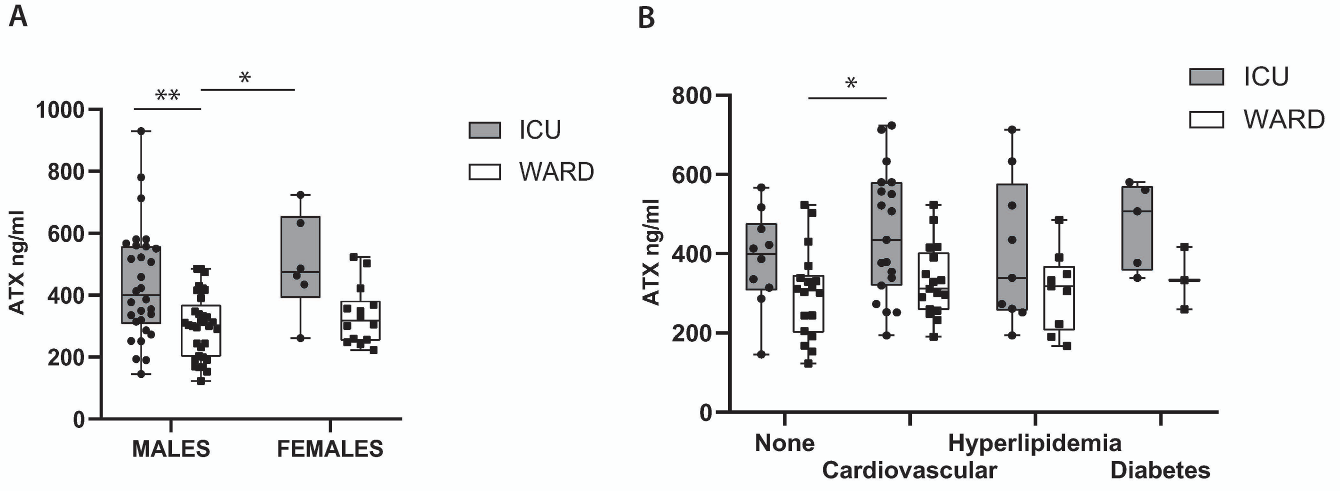


**Figure S1. Serum ATX protein levels in COVID-19 patients hospitalized in the ICU do not corelate with gender or comorbidities.** ATX protein levels were measured, with a commercial ELISA kit, in the sera of COVID-19 patients hospitalized (without Dex treatment) in the COVID-19 Ward (n=47) or the ICU (n=37) of Evaggelismos hospital. No significant differences in serum ATX levels were detected in COVID-19 patients, from either ICU/WARD, between **(A)** genders or **(B)** upon the presence of different comorbidities. Statistical significance, given the normal distribution of values, was assessed with 2-way ANOVA followed by Bonferroni post hoc correction. $^{*,**}$ denote p<0,001, 00001 respectively.

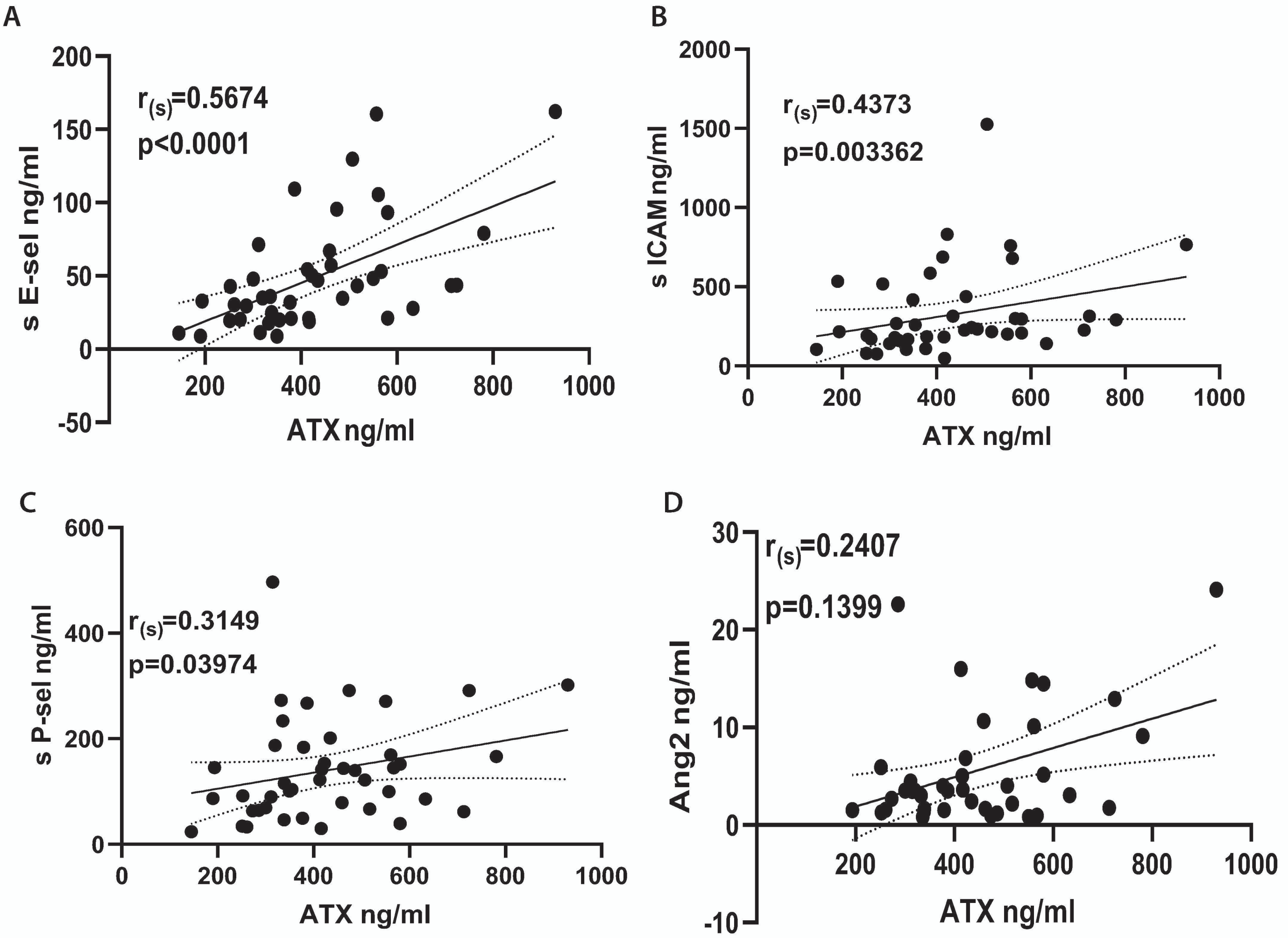


**Figure S2. ATX serum levels of ICU COVID-19 patients corelate with endothelial dysfunction biomarkers.** ATX serum levels in a subset of patients, reported at Figure 2, were corelated with the protein levels of **(A)** E-sel, **(B)** sICAM, **(C)** sP-sel and **(D)** ANG2, as measured and reported previously for the same patient samples. Statistical significance, indicated at each panel, was assessed with Spearman correlation ($r_s$).

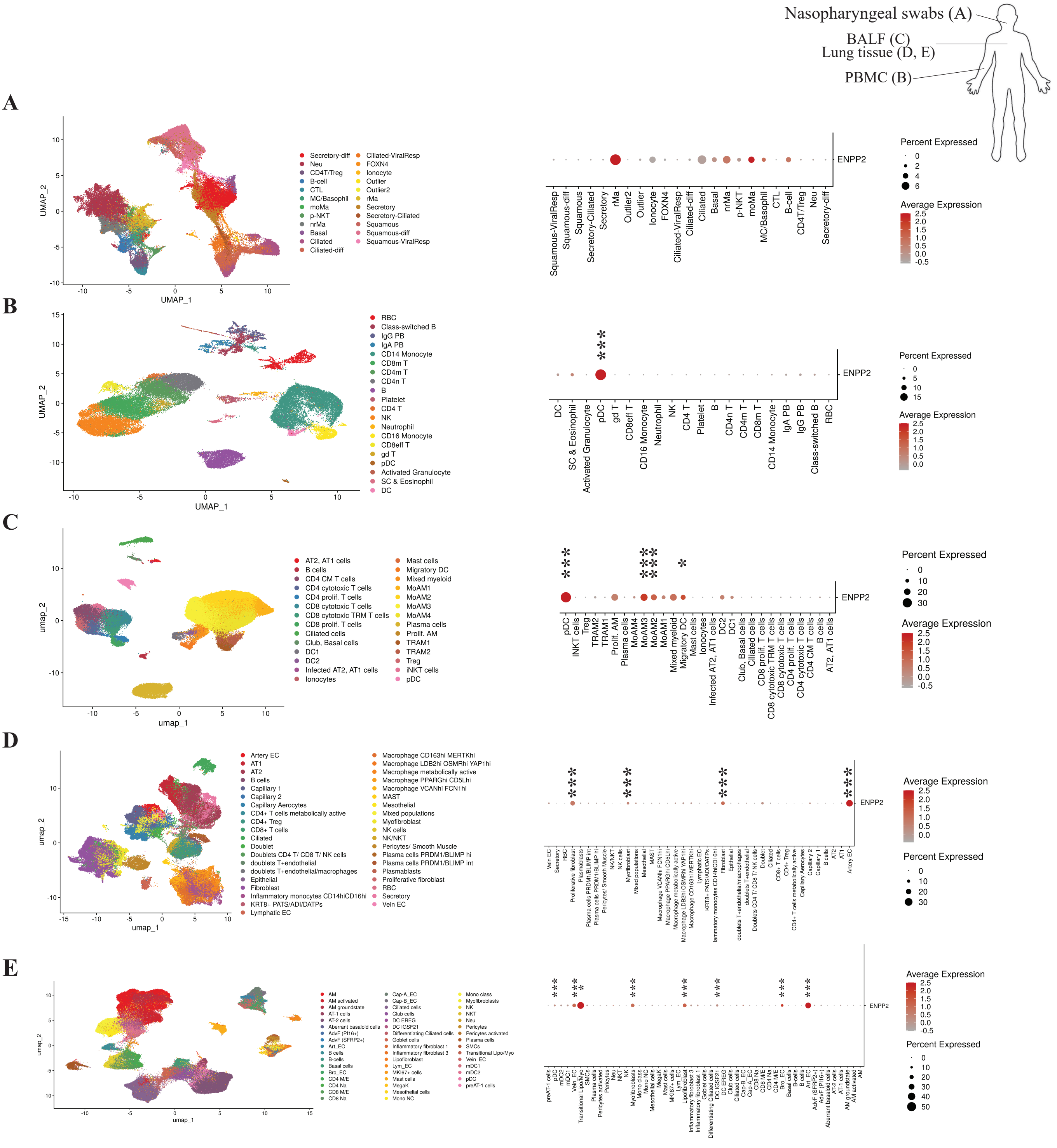


**Figure S3. The *ENPP2* expression landscape in COVID-19 and ILDs.** *ENPP2* expression has been assessed in four COVID-19 datasets **(A-D)**, each representing a different sampling site, as depicted, complementing the datasets shown in Figure 4. **(E)** A similar analysis was performed on an Interstitial Lung Diseases (ILD) scRNAseq dataset (Table S1). UMAP plots (on the left) depict the cellular composition of these sites, while dot plots (on the right) the expression pattern of *ENPP2* in the detected cell types. Marker genes, denoted by stars, were detected using using a Wilcoxon rank sum test; FC>1.2, Bonferroni corrected p<0.05; ***denotes p<0.01(PMIDs: A. 33361824; B. 32514174; C. 33429418; D. 33915569; E. 33650774).

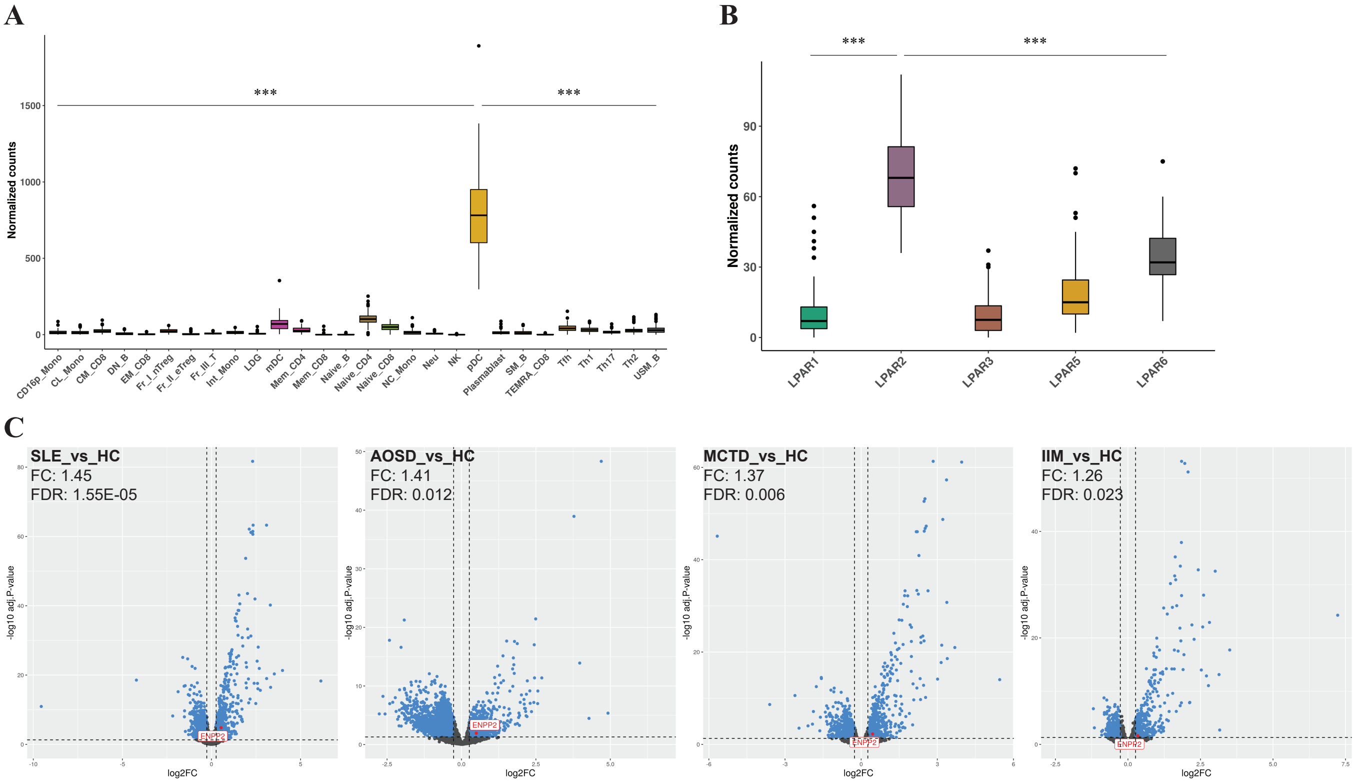


**Figure S4.** ***ENPP2*** **is differentially expressed in peripheral plasmacytoid dendritic cells (pDCs) during inflammatory diseases. (A).** During steady state conditions *ENPP2* is expressed higher in pDCs relative to other immune cell types. Boxplots depicting EDASeq normalized counts of *ENPP2* in various immune cells of healthy individuals with pairwise differential expression analysis between each cell type pDCs; ***denotes FC>1.2; FDR corrected p< 0.01. **(B).** *LPAR2* is the main receptor expressed by healthy pDCs. Boxplots depict the EDASeq normalized counts of the different LPA receptors. Statistical significance was assessed using Kruskal-Wallis and Dunn post-hoc test; ***denotes Benjamini-Hochberg adjusted p<0.01. **(C).** pDCs originating from four immune-mediated disorders express higher levels of *ENPP2* mRNA, as compared to their healthy counterparts; FC>1.2 and FDR corrected p<0.05. (PMID: 33930287) *HC: Healthy Control; SLE: Systemic Lupus Erythematosus; AOSD: Adult-Onset Still's Disease; MCTD: Mixed Connective Tissue Disease; IIM: Idiopathic Inflammatory Myopathy.*
